# Omicron outbreak at a private gathering in the Faroe Islands, infecting 21 of 33 triple-vaccinated healthcare workers

**DOI:** 10.1101/2021.12.22.21268021

**Authors:** Gunnhild Helmsdal, Olga K Hansen, Lars F Møller, Debes H Christiansen, Maria Skaalum Petersen, Marnar F Kristiansen

## Abstract

There are concerns that the SARS-CoV-2 Omicron variant evades immune responses due to unusually high numbers of mutations on the spike protein. Here we report a super-spreading event of Omicron infections amongst triple-vaccinated healthcare workers, infecting 21 of 33 attending a private gathering in the Faroe Islands.

---

South African researchers were the first to report the B.1.1.529 variant of the Severe Acute Respiratory Syndrome Coronavirus 2 (SARS-CoV-2) on November 24 [1]. Two days later, WHO named the variant Omicron and classified it as a variant of concern (VOC) [2].

The Omicron variant carries from 26 to 32 mutations on the spike protein, the main antigenic target of antibodies generated by infection or vaccination [3,4]. Early reports suggest that Omicron displays higher transmissibility and immune escape potential than earlier variants, while a neutralization study has shown lower neutralization activity for previously infected and vaccinated against the Omicron variant [5].

Although many countries worldwide have enacted travel restrictions to restrict the transmission of the Omicron variant, the variant has managed to spread quickly within Europe and beyond during the first weeks of December 2021. The fast spread has led to the expectation that the Omicron variant will become the dominating variant within a short time, and surveillance of the severity and transmissibility of the Omicron variant is critical in the following weeks and months [6].

The Faroe Islands, a self-governing group of islands located between Iceland and Norway, have been relatively successful in containing the COVID-19 pandemic [7,8]. On December 30, 2020, the first Faroese received the BNT162b2 vaccine (Comirnaty; BioNTech, Mainz, Germany), the only vaccine used in the Faroe Islands, and since a high proportion of the population has been vaccinated. As of December 8, 74.6% of the population has been vaccinated two times, and 13.6% three times [9]. From March 2020 to September 2021, the Faroe Islands had registered only 1,001 cases of COVID-19 (1,867 per 100,000) and two deaths (3.73 per 100,000). However, after loosening restrictions and the introduction of the more contagious Delta variant, a large outbreak has plagued the islands, with more than 3.300 cases and 11 deaths registered during the last two and a half months [9].

This paper reports a super-spreading event where 21 of 33 healthcare workers were infected with the Omicron variant after attending a social gathering in early December 2021, even though all infected participants had been vaccinated three times and had a recent negative test.

## Methods

The Chief Medical Officer’s office performs contact tracing in the Faroe Islands by interviewing all SARS-CoV-2 positive individuals and identifying their close contacts and their vaccination status [7]. Usually, only SARS-CoV-2 naive contacts are asked to quarantine. However, since the emergence of Omicron, all contacts to suspected Omicron positive individuals are asked to quarantine for seven days. They are also asked to get a PCR test immediately and on days 4 and 6, regardless of vaccination status. All positive individuals are required to isolate.

The Faroe Islands have performed among the highest numbers of COVID-19 tests per capita globally. Throat swabs are offered freely at several testing centers set up by the government, after which PCR analysis is performed at one of two centralized laboratories, with test results usually delivered within 12-24 hours. All cases connected to foreign travel, or where the Omicron variant is suspected, are genome sequenced by targeted sequencing, where parts of the spike protein are sequenced. This method is faster than performing whole genome sequencing on the virus, allowing for more cases to be screened.

All the positive individuals from the gathering agreed to participate in this study. They were interviewed by staff at the Chief Medical Officer’s office twice. The first time was shortly after the positive test, where they recorded the date of symptom debut, vaccination status, and personal characteristics. The second interview was performed 12-14 days after the exposure filling out a questionnaire, recording self-reported symptoms and severity, comorbidities, and medication.

## Results

During early December 2021, 33 persons attended a private gathering. Several participants noticed symptoms during the following days and performed a PCR test, which was positive. The other participants subsequently also performed PCR tests, resulting in 21 of 33 participants testing positive, corresponding to an attack rate of 63.6%. The unusually high attack rate led the Chief Medical Officer to request genome sequencing of the virus, identifying the first Omicron variant in the Faroes on December 8. So far, 13 samples from the gathering, and an additional four from the extended transmission chain, have been verified as the Omicron variant through targeted sequencing. The remaining cases are also assumed to be the Omicron variant. It has not been possible to definitively identify the index case initiating this transmission chain, but presumably, the variant has been imported from abroad.

All infected participants were fully vaccinated with the mRNA vaccine BNT162b2 (Comirnaty; BioNTech, Mainz, Germany) and had received a third booster dose within the last one and a half month, and none had a history of previous SARS-CoV-2 infection. One of the infected had received their third vaccine within days of the gathering, while the rest had received their vaccine at least three weeks before. The participants were unmasked during the gathering. The characteristics of the infected participants are displayed in Table 1. All infected participants had a negative test taken within 36 hours before the gathering. Most had performed a PCR test, while five had taken a lateral flow test. All the infected individuals experienced symptoms. The most common symptoms were muscle and joint pain, fatigue, and fever, while the least common symptoms were loss of taste and smell. No one was admitted to the hospital. We do not have individual-level information on the uninfected participants. However, we know that all had received at least two vaccines and had a negative test no more than 36 hours before.

**Table 1:** Personal characteristics and symptoms of Omicron infected individuals (n=21) from a gathering in the Faroe Islands

| Personal characteristics and symptoms |  |  |  |  |
| --- | --- | --- | --- | --- |
| Age, median | 45 years |  |  |  |
| Date of 1<sup>st</sup> vaccination* | 04-01-2021 to 15-01-2021 |  |  |  |
| Date of 2<sup>nd</sup> vaccination* | 05-02-2021 to 04-03-2021 |  |  |  |
| Date of 3<sup>rd</sup> vaccination* | 28-10-2021 to 02-12-2021 |  |  |  |
| Any comorbidity, n (%) | 4 (19%) |  |  |  |
| Any medication, n (%) | 3 (14%) |  |  |  |
|  | Symptoms (%) | Mild symptoms (%) | Moderate symptoms (%) | Severe symptoms (%) |
| Asymptomatic | 0 (0%) | 0 (0%) | 0 (0%) | 0 (0%) |
| Fever | 13 (62%) | 9 (69%) | 3 (23%) | 1 (8%) |
| Headache | 11 (52%) | 4 (36%) | 5 (45%) | 2 (18%) |
| Dry cough | 13 (62%) | 9 (69%) | 3 (23%) | 1 (8%) |
| Wet cough | 8 (38%) | 6 (75%) | 2 (25%) | 0 (%) |
| Dyspnea | 8 (38%) | 7 (88%) | 1 (13%) | 0 (%) |
| Loss of smell | 4 (19%) | 1 (25%) | 3 (75%) | 0 (%) |
| Loss of taste | 5 (24%) | 2 (40%) | 3 (60%) | 0 (%) |
| Fatigue | 15 (71%) | 4 (27%) | 7 (47%) | 4 (27%) |
| Rhinorrhea | 12 (57%) | 5 (42%) | 3 (25%) | 4 (33%) |
| Sinusitis symptoms | 6 (29%) | 3 (50%) | 1 (17%) | 2 (33%) |
| Throat pain | 10 (48%) | 7 (70%) | 2 (20%) | 1 (10%) |
| Myalgia | 13 (62%) | 7 (54%) | 4 (31%) | 2 (15%) |
| Arthralgia | 11 (52%) | 7 (64%) | 3 (27%) | 1 (9%) |
| Chest pain | 4 (19%) | 2 (50%) | 2 (50%) | 0 (%) |

\* All infected individuals had received three vaccinations

If we assume that the exposure to SARS-CoV-2 was on the evening of the gathering, the time to symptom onset was short, ranging from 2 to 6 days, with a mean incubation period of 3.24 days (95% CI 2.87-3.60). Time to resolution of symptoms varied, and at the end of follow-up, five individuals still reported symptoms, while the rest reported symptoms lasting 1 to 9 days.

## Discussion

This report demonstrates that the Omicron variant can lead to super-spreading events even in triple-vaccinated people. All the reported cases were symptomatic. None were admitted to the hospital. There is an urgent need for research into the Omicron variant, as with the current growth rates, it is anticipated to dominate the spread within a short time [10].

The primary spread of SARS-CoV-2 is through human-to-human transmission, either through respiratory droplets or aerosols. Factors such as indoor settings, poor ventilation, loud talking, laughing, and singing will increase the likely spread of the virus [11]. This study shows the ongoing importance of social distancing and the avoidance of larger festive gatherings during the pandemic in preventing possible super-spreading events. The active transmission chain described in this study was seemingly stopped after about 70 cases, demonstrating that through effective contact tracing, an Omicron transmission chain can be contained.

There is still limited clinical data available on the Omicron variant. However, Brandal et al. have reported a similar outbreak to the one reported in this paper in Norway, with an high attack rate of 74%, and similar symptoms to the cases reported here [12]. Furthermore, Espenhain et al. have published an early report on the first 785 Omicron cases in Denmark. They highlight the rapid spread of this variant and the ability to induce super-spreading events and that a high proportion of the infected was fully vaccinated. Since there is a lag time from infection to hospitalization and death, and this variant has only recently been discovered, sufficient data is not available to make conclusions on the severity of this variant yet [13].

Our findings indicate that the Omicron variant displays potent immune-escape properties and that even individuals recently boosted are at risk of getting infected. The incubation period of Omicron was short in this study. If the incubation period for Omicron is shorter than for previous variants, this can potentially partly explain the increased infection in individuals with some immunity.

There are several limitations to the observations in this report. Of note, the fact that many were infected in this gathering does not negate the possibility that the COVID-19 vaccines can prevent infections. It is not unlikely that we would have observed a higher attack rate, if the infected participants had not been triple-vaccinated. Also, the setting of a social gathering, with close contact between participants, does not necessarily generalize to other settings, since the amount of virus in the air will be higher in such social settings, increasing the risk of infection even among triple-vaccinated individuals. It is not possible to determine hospitalization rate or death rates from this small study. We do not yet know the risk of developing Long Covid after an Omicron infection. Even if the cases in this study primarily experienced relatively mild disease, all the reported cases have had previous immunity through vaccination.

It is notable, that all the infected cases experienced symptoms, and that especially loss of taste and smell seem to be less common in these cases, compared with previous outbreaks. It is likely that vaccination also protects against severe disease with the Omicron variant, even if protection against infection has waned to some degree, still underlining the importance of vaccination. Of note, the findings might not generalize to SARS-CoV-2 naive individuals, and for this reason, further research in Omicron amongst SARS-CoV-2 naive individuals is needed.

## Data Availability

All data produced in the present study are available upon reasonable request to the authors

## Author contributions

GH and MFK wrote the manuscript. OKH and LFM were responsible for contact tracing and interviewing the participants. DHC was responsible for the sequencing of the viral genomes. GH, OKH, LFM, DC, MSP and MFK contributed to the interpretation of data and read, edited, and approved the final manuscript.

## Acknowledgements

We would like to thank the staff at the Chief Medical Officer’s office and at the Faroese Food and Veterinary Authority, for their tireless work in connection with the COVID-19 pandemic.

## Ethical statement

The Faroese Ethical Committee ruled that this project belonged to the category of registry research and was not obliged to seek approval from the committee.

